# Longitudinal Analysis of Imaging Features of the Aortic Wall in Patients Diagnosed with an Aortic Dissection

**DOI:** 10.1101/2025.04.21.25326154

**Authors:** Grace H. Miner, Rami O. Tadros, Peter Faries, Michael L. Marin

## Abstract

**Background:** The presence of an aortic intimal flap is often identified following an acute aortic dissection (AD). However, the changes that take place in the dissected aorta have not been well characterized in late stage medically managed dissections in the descending thoracic aorta. The objective of this study is to evaluate the temporal changes observed on CT scans that take place in the aorta following an AD.

**Methods:** Patients were identified using ICD10 codes (I71.0) for aortic dissection. Inclusion criteria included persistent dissection distal to the left subclavian and CTA performed at the time of diagnosis and serially over a minimum of two years. Imaging features assessed include other affected arteries, wall morphology, (aortic wall thickness, calcification, irregularity), maximum diameter, and false lumen thrombosis.

**Results:** The mean age of the 44 aortic dissection patients meeting inclusion criteria was 59 years. The presence of luminal projections and ulcer-like-projections both significantly increased in the descending thoracic aorta over time (p=0.02 and p=0.003, respectively). The thoracic aortic wall appeared significantly more calcified at the time of the latest follow up scan (p=0.015). Additionally, increased frequency of descending thoracic and infrarenal aortic aneurysms were identified on CT scans in patients with long-term follow up (p=0.032 and p=0.018, respectively).

**Conclusion:** Following an AD, the aortic wall undergoes significant changes over time including dilation, calcification, and wall irregularity. Additionally, resolution of the traditional double barrel appearance of an AD can occur and obscure the imaging history of the dissection event.

## Introduction

An aortic dissection (AD) begins as an intimal tear which allows blood to penetrate between the layers of the aortic wall resulting in development of two lumens of blood flow (Figure 1). AD is relatively uncommon with an incidence of 3 per 100,000[1, 2]. Risk factors for developing AD include uncontrolled hypertension, atherosclerosis, smoking, and genetic disorders such as Marfan syndrome or Familial Thoracic Aortic Aneurysm and Dissection (FTAAD) syndrome [3]. AD is more common in men between the ages 50 and 70, and generally presents with severe chest pain that can simulate an acute myocardial infarction [4]. On initial presentation the diagnosis of AD is often confirmed with computed tomography (CT)[5]. Correct and early diagnosis of AD is essential as complications of AD can have a high morbidity and mortality [6].

**Figure 1.**
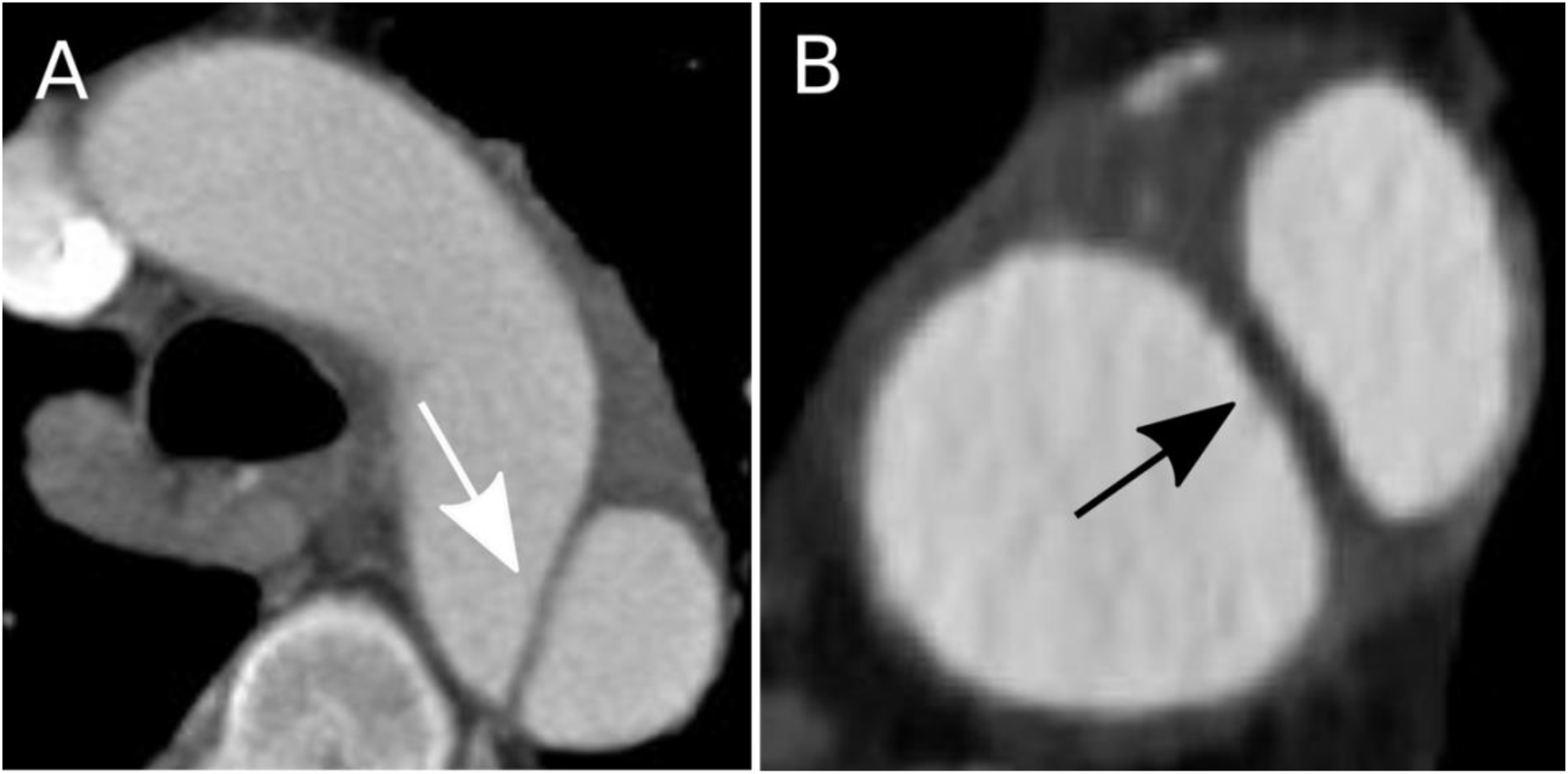
Axial view of computed tomography scans from two patients who were diagnosed with an aortic dissection. A) Aortic dissection flap (white arrow) just distal to the left subclavian artery in a patient with a type B dissection, B) Aortic dissection flap (black arrow) in the descending thoracic aorta of a patient with a type B dissection.

Dissections that begin in the ascending aorta (Debakey type I and II or Stanford type A) are considered emergent and require immediate surgical repair [7]. Conversely, AD originating in the descending thoracic aorta at or distal to the left subclavian artery (Debakey type III or Stanford type B) are less frequently associated with an acute surgical emergency and are often observed in the acute setting. If a patient with a type B dissection develops complications such as malperfusion, the dissection is repaired with a graft or endograft otherwise the patient is managed medically. Management includes blood pressure and pain control, with further monitoring for malperfusion or rupture of visceral, renal, or lower extremity vessels [8–12]. Type B dissections often remain stable for many years and patients receive routine annual CT scans to evaluate their dissections for any changes or remodeling [13].

Meaningful progress has been made for the surgical and endovascular treatment of AD, however questions remain regarding the natural history of the aorta after an AD are still unresolved. Progressive aortic degradation following an AD is a well observed finding in both sporadic, familial, and syndromic AD patients [14]. Approximately 40% of patients who receive initial medical management of their AD experience aneurysmal degeneration on follow-up studies [15]. Aneurysmal degeneration following AD may be secondary to increased degradation of elastin in the aortic wall with fibrosis over time [16, 17]. Histology studies have provided important insight into the structure and composition of the aortic wall following interventions to repair the AD, but have provided little information about the stages or events that occurs as the aorta resolves or heals from an acute AD. Comprehensive assessments of sequential CT scans of chronic medically managed AD patients are limited. Furthermore, assessment of imaging variables, and data collection methods have not been standardized. Without specific methods and standard definitions, comparisons between study results are difficult. The objective of this study is to evaluate the natural history of aortic dissection in serial CTAs and to establish a clearer understanding of the temporal changes that occur in the aorta following an initial dissection.

## Methods

### Patient Selection and Comorbidities

Patients diagnosed with aortic dissections were identified from the medical records using ICD10 codes resulting in a cohort of 659 patients. Forty-four patients were identified with dissections in the descending thoracic aorta who had received CT imaging at initial presentation, one year, and a minimum of two years after initial presentation. Patients with medically managed type B dissections and patients who underwent emergency ascending aortic repair who had persistent descending aortic dissections distal to the left subclavian artery following the intervention were included in this study. Patient demographics including age, sex, smoking history, and comorbidities such as hypercholesterolemia, hypertension, lung disease, and coronary artery disease were collected from the electronic medical records and included in this analysis. The Institutional Review Board (IRB) of the Mount Sinai Health System gave ethical approval for this work (IRB#: STUDY-11-00416) and all study activities were conducted in compliance with institutional guidelines.

### Imaging

All patients included in this study received serial high-resolution CT imaging of their chest, abdomen, and pelvis. Vitrea® Software was then used to reconstruct the multi-slice scans and used to confirm the presence and location of the dissection and evaluate imaging variables. The aorta was evaluated across multiple zones and seven imaging features of the aortic wall and lumen were assessed. Aortic diameter, aortic wall thickness, wall calcification, ulcer-like projections (ULPs), luminal penetrations, septal calcification, and level of lumen thrombus were assessed in the ascending aorta, aortic arch, and descending aorta (Figure 2). The maximum thickness of the aortic wall was assessed on axial scans and the greatest wall thickness in an area without thrombus was recorded. The largest continuous calcified lesion was measured on axial scan. ULPs were defined as small contrast filled ulcers originating from the lumen and extending into the aortic wall. Luminal penetrations were defined as any aortic wall material extending from the wall of the aorta and projecting into the lumen. If a scan showed the presence of both ULPs and luminal penetrations, it was classified as having aortic wall irregularity. Calcification of the dissection flap or residual dissection flaps was termed septal calcification. Thrombus was divided into three categories: complete thrombus, partial thrombus (any thrombus in the false lumen), or absence of thrombus in the false lumen. In addition to measurements of the wall and lumen, aneurysmal degeneration was assessed in both the descending thoracic and abdominal aorta.

**Figure 2.**
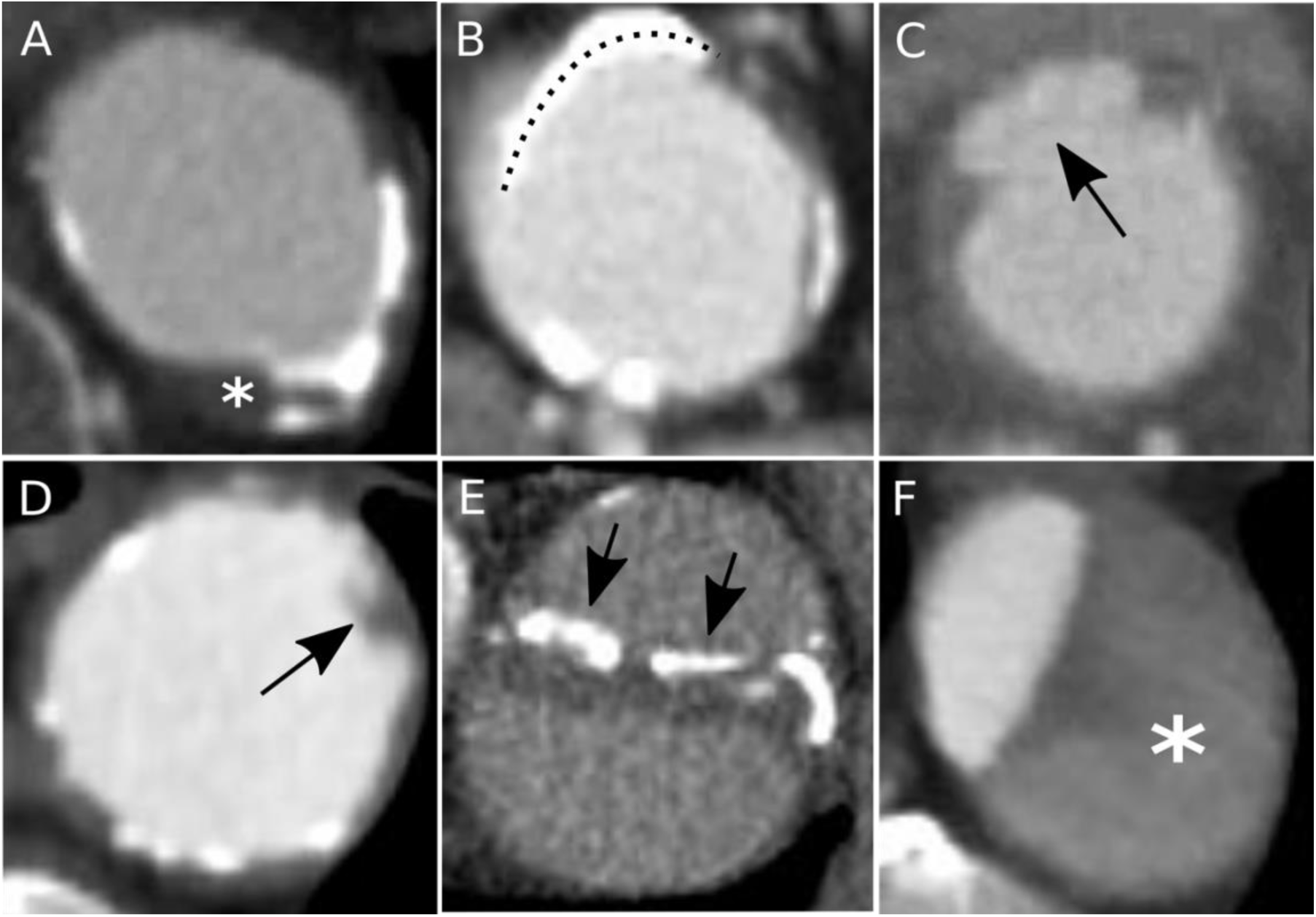
CT scans of patients with acute and chronic aortic dissections. A) Maximum aortic wall thickness in an area without thrombus (asterisk) in a patient with a chronic type B aortic dissection. B) Measurement of the circumferential length of calcium (dotted line) in a patient with a type B aortic dissection. C) Ulcer-like-projection (black arrow) in a patient with a chronic aortic dissection. D) Luminal projection (black arrow) in a patient with a chronic type B aortic dissection. E) Calcified septum (black arrows) in a patient with a late stage chronic type B aortic dissection. F) Thrombosed false lumen (asterisk) in a patient with a type B aortic dissection.

**Figure 3.**
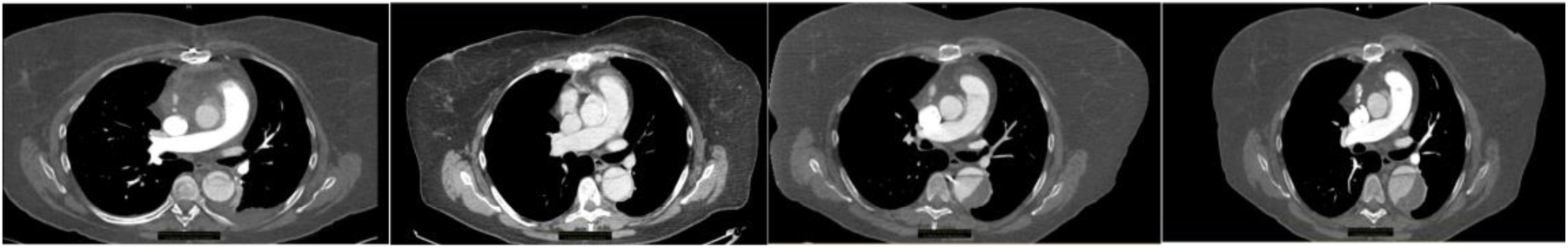
Changes in the aortic wall following an aortic dissection. The location of the dissection flap can change over time with enlargement of the false lumen and increasing amounts of thrombosis.

Aneurysms were defined as 1.5 times the diameter of the aorta superior to the area of enlargement. The extension of an aortic dissection into the abdominal aorta and iliac vessels was also noted.

### Statistics

The categorical data was analyzed using a chi-square tests or Fisher’s exact test, and expressed as either proportions or percentages. Shapiro tests were used to determine normality of the data. Normally distributed continuous data was expressed as mean ± standard deviation and was analyzed using analysis of variance. Nonparametric data was expressed as median and interquartile range and analyzed using the Kruskal-Wallis tests. A p value of < 0.05 was considered statistically significant. Patients with any missing data points were excluded rather than defaulting to negative. Following univariate statistics, any variable reaching a statistical significance p-value of 0.05 was included in the multivariate analysis. Statistics were performed using R software and associated libraries.

## Results

### Demographics and comorbidities

A total of 44 patients were identified who met inclusion criteria including CT scans at onset of AD, at 1 year, and 2 years or more following an acute dissection and had not undergone surgical repair. The CT scans and medical records of these 44 patients were reviewed. The mean age of the patients with AD was 59 years ± 10.3 at the time of presentation. Of the 44 AD patients, 29 (65.9%) were male and 15 (34.1%) were female. At the time of presentation of an acute dissection, 95.8% of patients were hypertensive (Table 1).

**Table 1.** Demographics of patients with aortic dissection.

|  | Overall |
| --- | --- |
| n | 44 |
| Age (mean (sd)) | 59.35 (10.28) |
| Male (%) | 29 (65.9) |
| Smoking (%) | 22 (50.0) |
| Coronary Artery Disease (%) | 24 (54.5) |
| Heart Failure (%) | 11 (25.0) |
| Hernia (%) | 12 (27.3) |
| Hypercholesterolemia (%) | 22 (50.0) |
| Hypertension (%) | 42 (95.5) |
| Kidney Disease (%) | 18 (40.9) |
| Lung Disease (%) | 8 (18.2) |
| Myocardial Infarction (%) | 4 (9.1) |

### AD time-course analysis

The 44 patients with AD were evaluated on initial presentation, 1 year, and ≥2 years after their initial diagnosis of AD on CT imaging. On initial CT imaging, 34 patients had DeBakey type I AD, 6 patients had DeBakey type IIIa, 4 patients had Debakey type IIIb. Of the 34 patients with a Debakey type I AD, five patients’ dissections extending into the abdominal aorta and one of the iliac. These five patients also had an additional dissection of an arch vessel, visceral vessel, or the internal or external iliac artery. Six patient’s aortic dissections appeared to fully resolve over the course of the study (p=0.04). Additionally, the number of Debakey type I dissections significantly decreased from 34 (77.3%) to 19 (43.2%) (p=0.004) and the number of Debakey type IIIb significantly increased from four (9.1%) to 11 (25%) (p=0.049) over the study period. Dissections in the descending thoracic aorta completely thrombosed the false lumen thrombosis of the dissection in 20.5% of the patients over the study period, p = 0.016 (Table 2).

**Table 2.** Computed tomographic analysis at the level of the descending thoracic and infrarenal aorta of patients with aortic dissections.

|  | Time=0 years | Time>1 years | Time=>2 years | P (univariate) | P value (multivariate) |
| --- | --- | --- | --- | --- | --- |
| n | 44 | 44 | 44 |  |  |
| Descending Thoracic aorta |  |  |  |  |  |
| Luminal projections (%) | 18 (40.9) | 25 (56.8) | 31 (70.5) | 0.016 | 0.393 |
| Ulcer-like-projections (%) | 13 (29.5) | 22 (50.0) | 29 (65.9) | 0.005 | 0.526 |
| Septal calcium (%) | 13 (29.5) | 9 (20.5) | 18 (40.9) | 0.121 |  |
| Aneurysm (%) | 20 (45.5) | 27 (61.4) | 32 (72.7) | 0.027 | 0.242 |
| Flap Thickness [median [IQR]] | 2.10 [1.80, 2.60] | 2.40 [2.20, 3.05] | 2.50 [2.08, 3.02] | 0.098 | 0.226 |
| Maximum Diameter [median [IQR]] | 36.55 [32.08, 41.15] | 39.10 [35.65, 43.42] | 40.30 [36.88, 45.65] | 0.139 | 0.880 |
| Wall Thickening [median [IQR]] | 4.25 [3.18, 6.55] | 3.85 [2.90, 6.50] | 3.60 [3.10, 4.32] | 0.418 |  |
| Wall Calcification [median [IQR]] | 0.00 [0.00, 9.60] | 0.00 [0.00, 9.03] | 5.85 [1.88, 12.20] | 0.017 | 0.585 |
| False Lumen Thrombosis (%) |  |  |  | 0.016 |  |
| 0.016 |  |  |  |  |  |
| No | 27 (61.4) | 26 (59.1) | 19 (43.2) |  |  |
| Partial | 17 (38.6) | 15 (34.1) | 16 (36.4) |  | 0.073 |
| Yes | 0 (0.0) | 3 (6.8) | 9 (20.5) |  | 0.994 |
| Infrarenal aorta |  |  |  |  |  |
| Luminal Projection (%) | 12 (27.3) | 19 (43.2) | 28 (63.6) | 0.003 | 0.679 |
| Ulcer-like-projections (%) | 10 (22.7) | 16 (36.4) | 24 (54.5) | 0.009 | 0.893 |
| Septal calcium (%) | 15 (34.1) | 14 (31.8) | 16 (36.4) | 0.904 |  |
| Aneurysm (%) | 13 (29.5) | 24 (54.5) | 25 (56.8) | 0.018 | 0.011 |
| Flap Thickness [median [IQR]] | 2.00 [1.60, 2.50] | 2.10 [1.80, 2.62] | 2.10 [1.85, 2.55] | 0.59 |  |
| Maximum Diameter [median [IQR]] | 27.90 [23.70, 31.75] | 30.10 [26.65, 34.62] | 32.20 [27.12, 38.30] | 0.071 |  |
| Wall Thickening [median [IQR]] | 3.10 [2.27, 4.50] | 2.75 [2.20, 3.35] | 3.00 [2.50, 4.03] | 0.323 |  |
| Wall Calcification [median [IQR]] | 5.25 [0.00, 11.12] | 6.80 [0.00, 11.80] | 7.70 [2.30, 13.77] | 0.216 |  |
| False Lumen Thrombosis (%) |  |  |  | 0.53 |  |
| 0.016 |  |  |  |  |  |
| No | 36 (81.8) | 34 (77.3) | 30 (68.2) |  |  |
| Partial | 6 (13.6) | 8 (18.2) | 9 (20.5) |  |  |
| Yes | 2 (4.5) | 2 (4.5) | 5 (11.4) |  |  |

Luminal projections and ULPs significantly increased over time in the descending thoracic aorta (p=0.02 and p=0.003 respectively). This trend was mirrored in the abdominal aorta, where the presence of both luminal projections and ULPs increased significantly (p=0.003 and p=0.009, respectively) (Table 2). The incidence of calcification of the septum between the two lumens was similar in the descending thoracic or abdominal aorta and was comparable between all three time-points. There was a significant increase in the wall calcification in the descending thoracic aorta over time. The median wall calcification increased from 0 to 5.85mm over the course of the study (p=0.015) (Table 2).

Importantly, 45.5% of the study cohort presented with thoracic aortic aneurysms at initial presentation. During the 2+ years of follow-up, the number of patients who developed a descending thoracic aneurysm in this population increased to 61.4% at 1 year and 72.7% after 2 years (p=0.32). Additionally, 29.5% of the study cohort were identified to have abdominal aortic aneurysms at the initial presentation. Over the study period, the number of the individuals with abdominal aortic aneurysms increased to 54.5% at one year and to 56.8% after two years (p=0.018) (Table 2). Of the 13 patients who presented with abdominal aortic aneurysms of any size, one patient had an abdominal aortic aneurysm measuring over 5cm. The presence of abdominal aortic aneurysm in the setting of a thoracic aortic dissection increased significantly over the 2 year study period (p=0.011).

## Discussion

Currently, comprehensive longitudinal imaging studies of AD are limited. Most studies focus on imaging features of the thoracic aorta in relation to surgical outcomes. Few imaging studies evaluate the natural history of the aorta over time from the descending thoracic aorta to the abdominal aorta. Over the course of the study, the aorta underwent changes including thrombosis, aortic enlargement, calcification, and changes in wall irregularity that exemplify the dynamic environment within the thoracic aorta following an AD (Figure 5). The selection of imaging features evaluated in patients with AD in this stud is consistent previous imaging studies[18]. ULPs, were defined as any focal, contrast filled pouch projecting into a thrombosed false lumen[19]. These ULPs have been observed in resolved dissections and in the absence of a previous diagnosis of AD are thought to be a potential precursor to AD[20]. The role of ULPs as a precursor to diffuse AD could not be confirmed in this study as all patients enrolled had been diagnosed with an AD as an inclusion criteria. However, ULPs were observed in patients with both active and resolved AD which suggests that ULPs may contribute to the progressive weakening of the aortic wall.

Increased diameter is another common metric for determining the progression of AD, as the aorta may enlarge over time degenerating into an aneurysm [15]. The percentage of patients with a descending thoracic aneurysm on presentation was over 40%, with a maximum aortic diameter of 45mm±12mm. More commonly, the descending thoracic aorta becomes aneurysmal following an AD. The International Registry of Acute Aortic Dissection (IRAD) observed descending thoracic aneurysm formation in 34.5% of uncomplicated type B aortic dissections at 3 years following AD, which increased to 73.3% at 5-year follow-up[13, 21]. While the rate of aneurysmal degeneration observed in the present study was comparable to the IRAD study, the maximum diameter of the descending thoracic aorta was slightly lower than previously reported [19, 22]. Greater aortic enlargement may be due to the high prevalence of multiple entry tears, which may be a risk factor for eventual aortic enlargement[23].

In addition to aortic enlargement, the presence of thrombus in the false lumen is a commonly reported metric in patients with AD (Figure 4). The degree of false lumen thrombosis is considered to be a predictor of poor surgical outcomes, eventual aortic enlargement, and ultimately mortality. Patients with partial thrombosis are considered at higher risk for rupture and have a higher mortality rate than patients who have fully thrombosed the false lumen of an AD [21, 24–26]. Partial thrombosis of the false lumen has also been linked to greater aortic enlargement and poor surgical outcomes[27].

**Figure 4.**
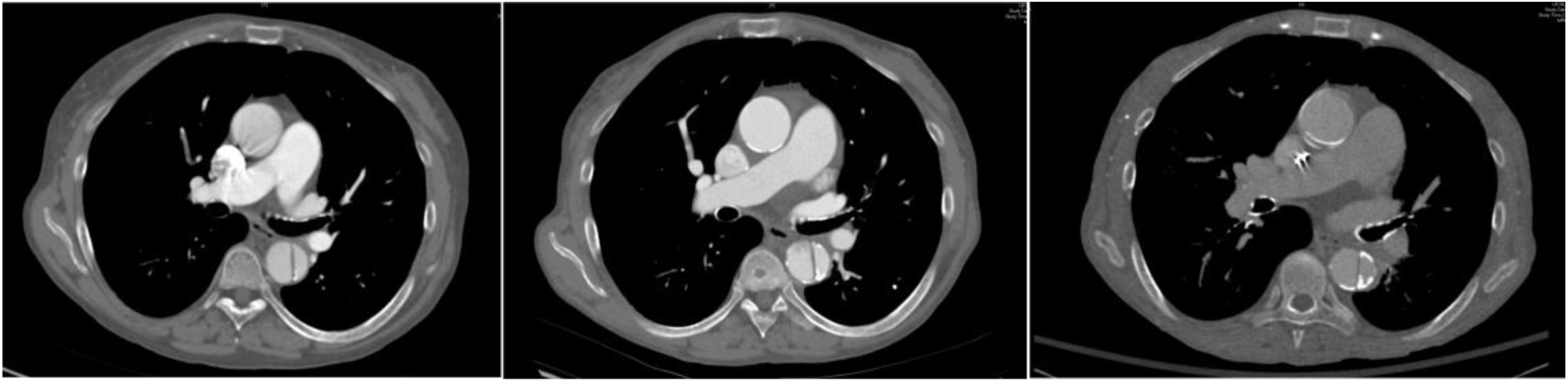
Thrombosis of the false lumen over time.

This may be due to the combination of a blockage of the exit tear and continued filling of the proximal tear, resulting in increased blood flow in the false lumen. The pressurized false lumen may then result in aneurysm formation, thinning the aortic wall, and an increased risk of rupture. This explanation is further supported by research showing that having greater than one entry tear results in poorer outcomes[28].

Surprisingly, an increase in calcification of the dissection flap over time was not observed. The observation of dissection flap calcification would be expected to suggest aging of the dissection flap, and thus progression of disease. However, dissection flap calcification was not uncommonly observed on initial presentation. Large calcified regions of the aortic wall were also observed, suggesting when the aorta dissects, the calcified lesions within the aortic wall tear free and become part of the septum (Figure 5).

**Figure 5.**
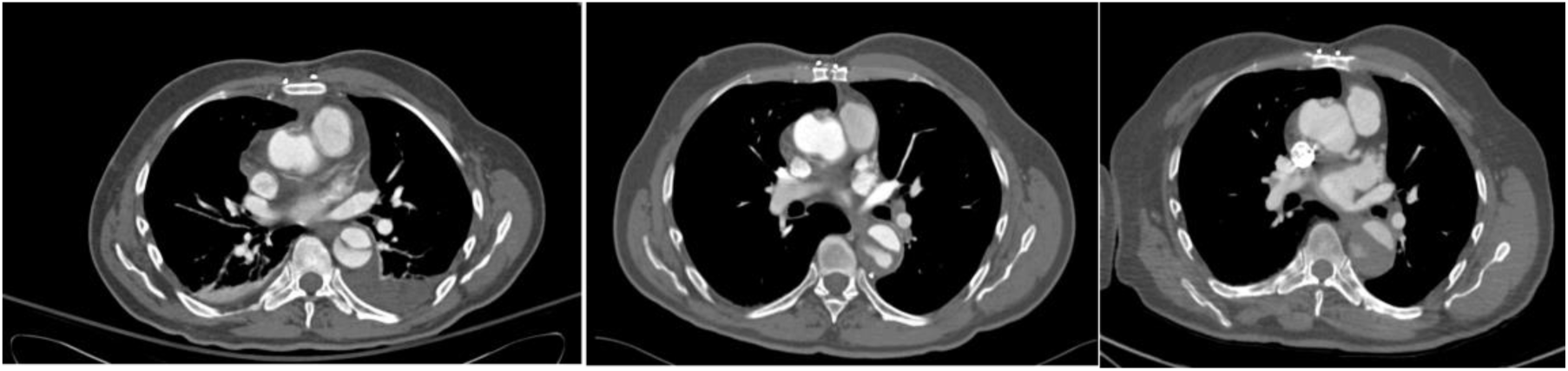
Increasing calcification of the aortic wall and dissection flap over time.

The results of this study showed that a subset of the dissections remodeled and resolved over time. Favorable remodeling was observed in 67% of patients with AD. The remodeling observed in patients with known DeBakey type I or III aortic dissections may be consistent with the features of diffuse aortic lumen irregularity seen in acute aortic syndrome (AAS). AAS includes patients with AD, penetrating aortic ulcers (PAU), intramural hematoma (IMH) and can present with chest pain and uncontrolled hypertension [29]. Previous studies suggest that AD, PAU, and IMH are a continuum of disease such that AD, PAU, and IMH are all interconvertible and can also be present at the same time [30]. Further research is required to determine the relationship between AD, PAU, and IMH and imaging features assessed in this study.

## Conclusion

The descending thoracic aorta undergoes significant changes from the acute to the chronic phase of AD. Over time, CT scans of patients exhibit increased thrombosis of the false lumen, increased wall calcification, and aneurysmal degeneration of the descending thoracic aorta. Additionally, the infrarenal segment of the aorta can exhibit significant aneurysmal degeneration. Full resolution of a subset of the aortic dissections can occur. Commensurate with false lumen thrombosis and remodeling, a subset of the dissections resolve such that the dissection flaps are no longer detectable. The results of this study show the aorta undergoes many changes following an aortic dissection. Understanding the natural history of AD and the changes within the aorta could influence management decisions and the outcome of surgical intervention.

## Data Availability

All data produced in the present study are available upon reasonable request to the authors

